# Survival and Factors Associated with Mortality Among Children with Congenital Heart Defects at the Uganda Heart Institute, Mulago National Referral Hospital

**DOI:** 10.64898/2026.04.30.26352117

**Authors:** Nassali Lovisa, Leonard Atuhaire, John Bosco Asiimwe, Dick Nsimbe

## Abstract

**Background:** Congenital heart defects (CHDs) contribute to approximately 220,000 childhood deaths globally each year, with most occurring in low- and middle-income countries. Despite advances in diagnosis and management, survival remains poor in sub-Saharan Africa due to delayed diagnosis and limited access to specialized care. In Uganda, an estimated 16,000 children are born with CHDs annually, many requiring urgent intervention. Despite this burden, evidence on survival and its determinants in Uganda remains limited. This study aimed to investigate socio-demographic and clinical factors associated with survival among children born with CHDs at the Uganda Heart Institute.

**Methods:** A retrospective cohort study was conducted using electronic patient records of children diagnosed with congenital heart defects at the Uganda Heart Institute between January 2014 and December 2018. Survival analysis was performed using the log-rank test to assess differences across groups, and the Cox proportional hazards regression model was used to identify independent predictors of mortality.

**Results:** Children residing in rural areas had a significantly higher hazard of death compared to those in urban areas (HR = 1.33; 95% CI: 1.06–1.66; p = 0.013). Underweight children had more than twice the hazard of death compared to those with normal BMI (HR = 2.07; 95% CI: 1.60–2.69; p < 0.001). Defect severity was significantly associated with survival, with moderate defects showing increased hazard relative to critical defects (HR = 1.98; 95% CI: 1.25–3.13; p = 0.004), while non-critical defects were not statistically significant. Timing of diagnosis was a strong predictor of mortality (HR = 1.67; 95% CI: 1.30–2.14; p < 0.001), indicating that delayed diagnosis increases the risk of death. Oxygen level was not significantly associated with survival.

**Conclusion:** Survival among children with congenital heart defects is significantly influenced by nutritional status, place of residence, defect severity, and timing of diagnosis. Underweight children, those from rural areas, and those diagnosed later have a higher risk of mortality. Early detection and improved nutritional support are essential to enhance survival outcomes. Strengthening early screening programs and improving access to timely diagnosis and specialized care, particularly in rural settings, are critical to reducing mortality.

## Background

Over the past three decades, substantial progress has been made in improving child survival globally, with under-five mortality declining by more than 50% between 1990 and 2019 (1,2). Despite these gains, congenital conditions particularly congenital heart defects (CHDs) have emerged as an important cause of childhood morbidity and mortality (3). CHDs are defined as structural abnormalities of the heart or great vessels present at birth and represent the most common congenital anomalies worldwide, affecting approximately 8–10 per 1,000 live births (4,5). Globally, CHDs account for an estimated 217,000–222,000 deaths annually, with nearly 90% of these deaths occurring in low- and middle-income countries (3).

The clinical spectrum of CHDs is broad, ranging from mild lesions that may remain asymptomatic to severe defects requiring immediate intervention (6). Critical congenital heart defects (CCHDs), which account for approximately 25% of all CHD cases, often require surgical or catheter-based intervention within the first year of life (7). In the absence of timely diagnosis and treatment, nearly one-third of children with severe CHDs die within the first month of life, and up to 50% do not survive beyond the first year (8,9). Although advances in diagnostic technologies, surgical techniques, and perioperative care have led to significant improvements in survival in high-income countries with survival rates exceeding 85% into adulthood, these gains have not been equitably realized in sub-Saharan Africa (10,11).

In Africa, an estimated 500,000 children are born with CHDs each year, with the highest burden in sub-Saharan Africa (11,12). However, more than 90% of these children lack access to appropriate cardiac care, resulting in high mortality rates (13). Studies across the region indicate that CHDs are a leading cause of pediatric heart failure, accounting for up to 30–40% of cardiac-related hospital admissions among children (14,15). In East Africa, efforts to establish pediatric cardiac services have been ongoing for the past two decades, often supported by international cardiac missions. Despite these initiatives, access to timely diagnosis and definitive treatment remains limited due to financial constraints, inadequate infrastructure, and shortages of specialized personnel (11,12).

In Uganda, CHDs continue to contribute significantly to childhood morbidity and mortality, posing a challenge to achieving Sustainable Development Goal 3 targets on child survival. According to reports from the Uganda Heart Institute, approximately 16,000 children are born with congenital heart defects annually in Uganda (16). A substantial proportion of these cases require urgent surgical or interventional care. The Uganda Heart Institute (UHI), located at Mulago National Referral Hospital, is the primary centre providing specialized pediatric cardiac services in the country (14). Since the introduction of open-heart surgery in 2007 and cardiac catheterization in 2012, the capacity for managing CHDs has steadily improved, with the number of procedures increasing annually and reported surgical mortality rates ranging between 0% and 4% (17). However, many children continue to present late with advanced disease, and survival outcomes remain suboptimal due to delays in diagnosis, limited access to care, and socioeconomic barriers.

Although several studies in Uganda have examined pediatric cardiovascular care, including service availability and quality of life among affected children (18–21), there remains limited evidence on survival patterns and the factors influencing it. there remains limited evidence on survival patterns and their determinants. Existing studies have largely focused on descriptive outcomes, with limited exploration of factors associated with mortality among children with CHDs. Therefore, this study aims to investigate the factors associated with survival among children diagnosed with congenital heart defects at the Uganda Heart Institute.

## Methods

The study was conducted using secondary data from the Uganda Heart Institute (UHI), located at Mulago National Referral Hospital in Kampala, Uganda, the country’s primary centre for specialized cardiac care. UHI provides diagnostic, medical, and surgical services for patients with cardiovascular diseases and maintains a comprehensive database of patient records. This retrospective cohort study analyzed data from children aged 0–18 years diagnosed with congenital heart defects (CHDs) between January 2014 and December 2018, with follow-up information available in the UHI database. Patient information, including demographic and clinical characteristics, is routinely recorded at admission and updated following diagnosis by cardiologists and cardiac surgeons, with most records stored in electronic format. Data extraction was conducted from these records, and only cases with complete registration and follow-up information were included in the analysis. Of approximately 13,000 patient records with heart conditions, 5,956 were identified as congenital heart defect cases with complete data, and these constituted the final analytical sample.

### Measures of Outcome

The dependent variable in this study was survival time, defined as the duration from birth to death or last follow-up, measured in months. The event of interest was death. Survival status was coded as 1 for death and 0 for censored observations. Children who were alive at the end of the study period or lost to follow-up were treated as right-censored observations, as the event of interest had not occurred during the observation period.

### Measures of explanatory Variables

The independent variables included demographic and clinical characteristics of the children. Gender was recorded as male or female. Place of residence was categorized as urban or rural based on the child’s current residence. Timing of diagnosis was measured as the age in months at which the child was diagnosed with a congenital heart defect (continuous). Anthropometric measures included height and weight (both continuous), from which Body Mass Index (BMI) was derived and categorized as normal or underweight. Oxygen level was categorized as normal or abnormal based on clinical assessment. Defect severity was determined by a cardiologist or cardiac surgeon and categorized as critical, moderate, or non-critical.

### Statistical Analysis

All data preparation and statistical analyses were conducted using Stata version 16. Descriptive statistics were used to summarize the demographic and clinical characteristics of the study population, with categorical variables presented as frequencies and percentages, and continuous variables summarized using appropriate measures of central tendency and dispersion. A survival analysis approach was employed to examine time to death among children with congenital heart defects. Survival functions were estimated using the Kaplan-Meier method, and differences between groups were assessed using the log-rank test. The Nelson-Aalen estimator was used to estimate cumulative hazard functions and to visualize differences in hazard across subgroups. At the multivariate level, the Cox proportional hazards regression model was fitted to assess the association between explanatory variables and time to death. Hazard ratios (HRs) with 95% confidence intervals were reported. Model diagnostics were conducted to evaluate the adequacy of the fitted model. Multicollinearity among explanatory variables was assessed using variance inflation factors (VIF), with no evidence of problematic collinearity observed. The proportional hazards assumption was assessed using Schoenfeld residuals. Model specification was evaluated using the linktest, where a statistically significant *hat* term and a non-significant *hat-squared* term indicated adequate model specification (22–25). Statistical significance was assessed at the 5% level.

## Results

Table 1 presents the distribution of child characteristics along with survival outcomes across selected socio-demographic and clinical variables. The table summarizes the frequency distribution, number of deaths, expected deaths, and log-rank test-statistics used to assess differences in survival functions across groups. The majority of the children included in the study were female (52.1%), while 47.9% were male. Most children (61.7%) resided in rural areas, compared to 38.3% from urban settings. With regard to nutritional status, the majority (76.6%) had normal body mass index (BMI), whereas 23.4% were underweight. In terms of oxygen saturation, 72.4% of the children had normal oxygen levels, while 27.6% had abnormal oxygen saturation. Regarding defect severity, most children (80.1%) had non-critical defects, 12.9% had critical defects, and 7.0% had moderate defects. For continuous characteristics, the median age at diagnosis was 14 months, with a minimum of 0 months and a maximum of 215 months. The mean height of the children was 91 cm (range: 5-109 cm), while the average weight was 12.68 kg (range: 3-87 kg).

Furthermore, Table 1 reveals that the log-rank test results indicate that there was no statistically significant difference in survival between male and female children (χ^2^ = 2.39, p = 0.122). However, place of residence was significantly associated with survival (χ^2^ = 4.97, p = 0.025), with children from rural areas showing differences in survival compared to their urban counterparts. Body mass index was strongly associated with survival outcomes (χ^2^ = 182.24, p < 0.001), indicating significant differences in survival between underweight and normal BMI children. Similarly, oxygen saturation level was significantly associated with survival (χ^2^ = 14.60, p < 0.001), suggesting that children with abnormal oxygen levels experienced different survival outcomes compared to those with normal levels. Defect severity was also significantly associated with survival (χ^2^ = 19.33, p = 0.002), indicating variation in survival probabilities across critical, moderate, and non-critical congenital heart defects. Overall, a total of 5,956 children were included in the study, with 315 deaths observed during the follow-up period. The Nelson–Aalen cumulative hazard estimates corresponding to these covariates are presented in Figure 1, showing a general increase in cumulative hazard over time across groups.

**Table 1.** Distribution of selected socio-demographic and clinical characteristics with survival outcomes among children with congenital heart defects.

| Covariates | N | Percentage | Observed deaths | Expected death | Log-rank<br>($\chi^2$)<br>p-value |
| --- | --- | --- | --- | --- | --- |
| **Gender** |  |  |  |  |  |
| Male | 2851 | 47.9 | 152 | 165.67 | 2.39 |
| Female | 3105 | 52.1 | 163 | 149.33 | (p = 0.122) |
| **Residence** |  |  |  |  |  |
| Urban | 2283 | 38.3 | 178 | 197.12 | 4.97 |
| Rural | 3628 | 61.7 | 137 | 117.88 | (p = 0.025) |
| **Body Mass Index** |  |  |  |  |  |
| BMI Normal | 4562 | 76.6 | 247 | 298.55 | 182.24 |
| BMI Underweight | 1394 | 23.4 | 68 | 16.45 | (p < 0.001) |
| **Oxygen level** |  |  |  |  |  |
| Normal | 4312 | 72.4 | 218 | 245.94 | 14.60 |
| Abnormal | 1644 | 27.6 | 97 | 69.06 | (p < 0.001) |
| **Defect Severity** |  |  |  |  |  |
| Critical | 769 | 12.9 | 38 | 56.32 |  |
| Moderate | 416 | 7.0 | 37 | 20.45 | 19.33 |
| Non-critical | 4771 | 80.1 | 242 | 238.23 | (p = 0.002) |
| **All** | **5956** | **100** | **315** | **315** |  |

**Fig.1.**
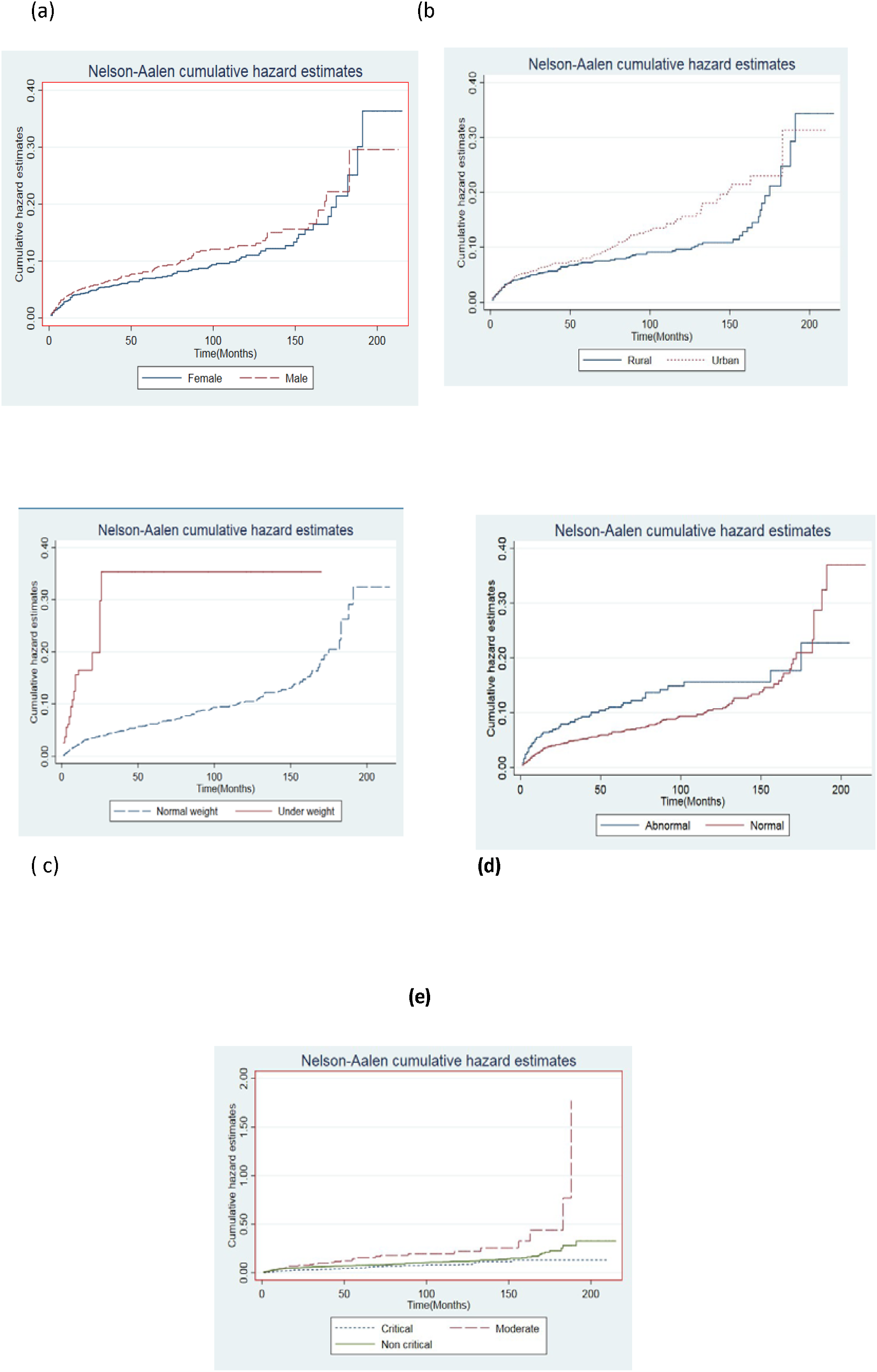
Nelson-Aalen cumulative hazard estimates: (a) Gender; (b) Residence; (c) Body mass index (BMI); (d) Oxygen level in the body; (e) Defect severity

### Factors Associated with Mortality Among Children with Congenital Heart Defects

To identify the independent factors associated with time to death among children with congenital heart defects, a multivariate Cox proportional hazards regression model was fitted using variables identified at the bivariate level. The factors included in the final model were place of residence, body mass index (BMI), oxygen saturation level, defect severity, and timing of diagnosis, as presented in Table 2. Results in Table 2 indicate that place of residence was significantly associated with mortality. Children residing in rural areas had a 32.8% higher hazard of death compared to those in urban areas (HR = 1.328; 95% CI: 1.06-1.66; p = 0.013), suggesting poorer survival outcomes among rural residents. Body mass index was also a strong predictor of mortality. Children who were underweight had more than twice the hazard of death compared to those with normal BMI (HR = 2.074; 95% CI: 1.60-2.69; p < 0.001), indicating substantially increased risk among undernourished children. Oxygen saturation level was not significantly associated with mortality (HR = 0.868; 95% CI: 0.68-1.11; p = 0.263), suggesting no evidence of a difference in survival between children with normal and abnormal oxygen levels after adjusting for other factors. Defect severity showed a significant association with mortality. Children with moderate defects had nearly twice the hazard of death compared to those with critical defects (HR = 1.979; 95% CI: 1.25-3.13; p = 0.004). However, non-critical defects were not significantly associated with mortality (HR = 1.307; 95% CI: 0.92-1.86; p = 0.134). Timing of diagnosis was a significant predictor of mortality. A unit increase in the timing of diagnosis was associated with a 67.0% increase in the hazard of death (HR = 1.670; 95% CI: 1.30–2.14; p < 0.001), indicating that delayed diagnosis substantially increases the risk of mortality among children with congenital heart defects. The proportional hazards assumption was assessed for the overall model and was not violated (p = 0.031), indicating that the Cox proportional hazards model was appropriate for the data. The linktest results further confirmed that the model was correctly specified. The *hat statistic* was statistically significant (p < 0.001), while the *hat-squared statistic* was not significant (p = 0.182), suggesting that the model was adequately specified and that no important variables were omitted.

**Table 2.**
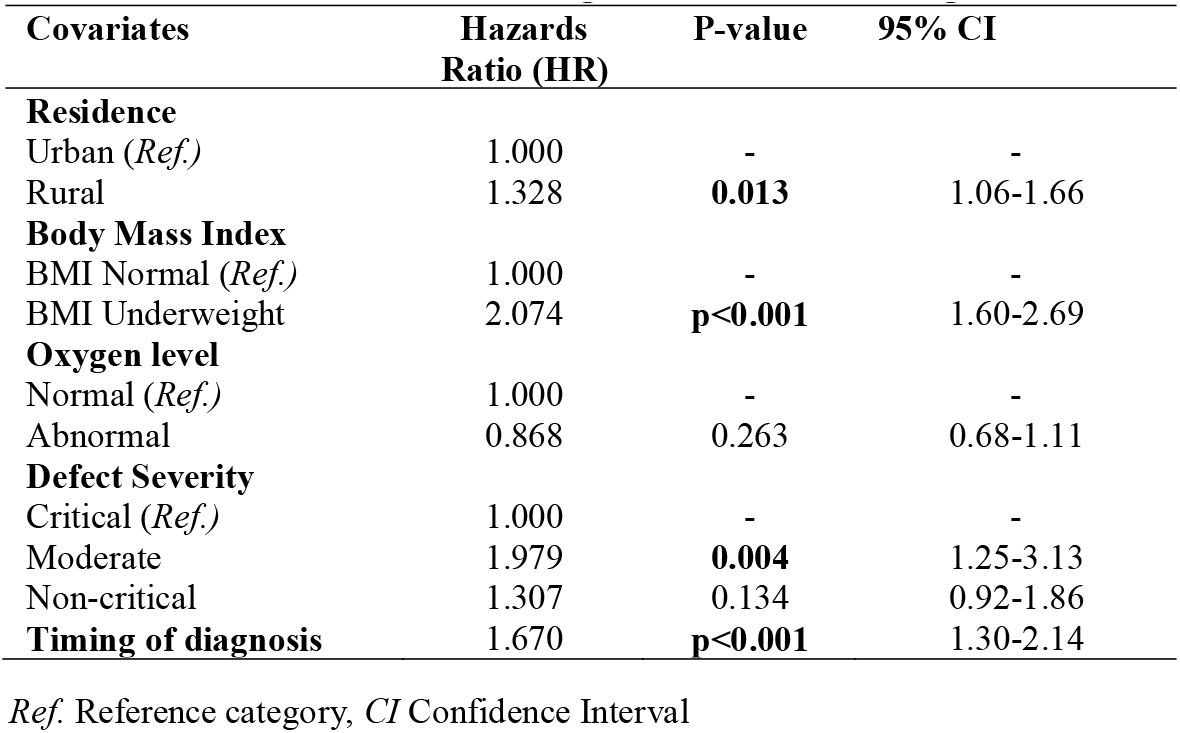
Cox proportional hazards model results linking socio-demographic and clinical factors to time to death among children with congenital heart defects.

## Discussion

This study provides empirical evidence on survival and its determinants among children with congenital heart defects in Uganda, addressing a critical gap in the limited literature on survival outcomes in this setting. The findings indicate that mortality is significantly influenced by socio-demographic and clinical factors, specifically, children diagnosed later, those who were underweight, and those residing in rural areas had a higher hazard of death compared to their counterparts. These findings highlight the critical role of early diagnosis, adequate nutritional status, and equitable access to specialized cardiac care in improving survival outcomes among children with congenital heart defects. The observed association between delayed diagnosis and increased mortality is consistent with existing literature, which emphasizes the importance of early detection in improving survival outcomes among children with CHDs. For instance, Eckersley et al. reported that late diagnosis is associated with excess mortality, while antenatal detection significantly reduces CHD-related deaths (26). Similarly, Animasahun et al. highlighted the need for improved awareness, early presentation, and strengthened referral systems to facilitate timely diagnosis and management (27). In low-resource settings such as Uganda, early diagnosis remains challenging, as some children with CHDs may appear clinically stable at birth and are discharged without detection, only to present later with severe complications requiring emergency care (17). This underscores the need for strengthened new-born screening programs, improved clinical vigilance, and enhanced referral pathways to ensure timely identification and management of congenital heart defects.

Place of residence was found to be a significant predictor of survival among children with congenital heart defects. Children residing in rural areas had a higher hazard of mortality compared to those in urban areas. This finding suggests that disparities in access to healthcare services may play a critical role in influencing survival outcomes. Children in rural settings often face challenges such as delayed access to specialized cardiac care, limited availability of diagnostic services, and weaker referral systems, which may contribute to late presentation and poorer outcomes (28). These findings are consistent with existing literature highlighting geographical inequalities in health outcomes. For instance, studies have shown that children living in rural areas are more likely to experience delays in diagnosis and treatment due to barriers such as long travel distances, limited healthcare infrastructure, and financial constraints (29). Langlois and colleagues also reported that variations in urban-rural classification may influence the detection and reporting of congenital heart defects, with milder cases being less frequently identified in rural settings (30). This suggests that differences in detection and access to care, rather than environmental factors alone, may explain the observed disparities in survival. Overall, the findings underscore the need to strengthen healthcare delivery systems in rural areas, improve referral pathways, and enhance access to specialized cardiac services to reduce mortality among children with congenital heart defects.

Body Mass Index (BMI) was found to be a significant predictor of survival among children with congenital heart defects. Underweight children had more than twice the hazard of death compared to those with normal BMI, indicating that poor nutritional status is strongly associated with increased mortality. Children with normal weight were therefore at a lower risk of death compared to their underweight counterparts. This finding is consistent with previous studies that have highlighted the critical role of nutritional status in the prognosis of children with CHDs. For instance, Majeed et al. reported that malnutrition and weight loss in children with congenital heart disease are associated with increased mortality (31). Similarly, Batte et al. found that malnutrition is highly prevalent among children with CHDs and is often accompanied by complications such as anaemia and heart failure, which further worsen clinical outcomes (18). The observed association may be explained by the increased metabolic demands and feeding difficulties commonly experienced by children with CHDs. These children often have poor feeding tolerance, increased energy expenditure, and recurrent infections, all of which contribute to growth failure and undernutrition. Consequently, malnutrition may exacerbate disease severity and reduce the child’s ability to withstand surgical or medical interventions. These findings highlight the need to integrate nutritional assessment and management into routine care for children with congenital heart defects. Early identification and treatment of malnutrition could play a critical role in improving survival outcomes in this population.

Defect severity was found to be a significant predictor of survival among children with congenital heart defects. Children with moderate defects had nearly twice the hazard of death compared to those with critical defects, while non-critical defects were not statistically significant. This finding may appear counterintuitive, as critical defects are generally associated with higher mortality; however, it may be explained by differences in clinical management and timing of intervention. Children with critical congenital heart defects are more likely to present with severe symptoms early in life, leading to prompt diagnosis and prioritization for specialized care and intervention (32). In contrast, children with moderate defects may present with less obvious symptoms, resulting in delayed diagnosis and treatment, which increases the risk of complications and mortality. These findings are consistent with previous studies suggesting that early detection and intensive management improve survival outcomes among children with severe CHDs. For example, Pace et al. reported that infants with critical CHDs experience high early mortality but are more likely to receive immediate and targeted care, which may improve survival among those who access treatment (33). Conversely, children with less severe but clinically significant defects may be overlooked or diagnosed later, contributing to poorer outcomes. Overall, these results highlight the importance of ensuring timely diagnosis and appropriate management across all levels of defect severity, not only for critically ill children but also for those with moderate conditions who remain at substantial risk.

In summary, this study utilized secondary data from 5,956 children diagnosed with congenital heart defects at the Uganda Heart Institute between 2014 and 2018, and survival analysis was conducted to examine time to death. The findings revealed that nutritional status, timing of diagnosis, place of residence, and defect severity were significant predictors of survival among children with congenital heart defects. Specifically, children who were underweight, diagnosed later, and those residing in rural areas had a higher risk of mortality. In contrast, oxygen level and non-critical defect severity were not significantly associated with survival outcomes.

There are some limitations that should be considered when interpreting these findings. The use of secondary data limited the ability to control for all potential confounding variables and restricted the analysis to variables available in the Uganda Heart Institute database. In addition, missing or incomplete information in some patient records may have affected the accuracy and reliability of the results. Furthermore, although age is an important factor in survival analysis, it was not included as an independent variable in this study because survival time was defined from birth, making age intrinsically accounted for within the time-to-event framework. Including age as a separate predictor could introduce collinearity and violate model assumptions, particularly since age at diagnosis was already captured through the timing of diagnosis variable. Additionally, this study was conducted at a single specialized cardiac centre, which may limit the generalizability of the findings to other healthcare settings, particularly those with limited access to specialized services. Future research should consider prospective study designs that allow for the inclusion of additional clinical, socioeconomic, and time-varying factors. In particular, incorporating age at different stages of disease progression, as well as longitudinal nutritional and clinical indicators, could provide deeper insights into survival patterns among children with congenital heart defects. Qualitative approaches may also help to better understand barriers to early diagnosis and access to care, especially in rural populations.

## Conclusion

This study identified key factors of survival among children with congenital heart defects at the Uganda Heart Institute, including delayed diagnosis, undernutrition, rural residence, and defect severity. These findings underscore the importance of early detection, improved nutritional support, and equitable access to specialized cardiac care. Strengthening new-born and early-life screening, improving referral systems, and ensuring timely diagnosis and treatment-particularly for children in rural areas are critical to reducing mortality. Integrating routine nutritional assessment and management into pediatric cardiac care may further improve clinical outcomes. Overall, targeted interventions addressing delays in diagnosis, healthcare access disparities, and nutritional challenges are essential to improve survival in this population.

## Data Availability

Data Availability: The datasets used and/or analyzed during the current study are not publicly available due to ethical and confidentiality restrictions but are available from the corresponding author on reasonable request and with permission from the Uganda Heart Institute.

## Abbreviations

AVSD: Atrioventricular Septal Defect
ASD: Atrial Septal Defect
BMI: Body Mass Index
CHD: Congenital Heart Defects
COA: Coarctation of the Aorta
CCHD: Critical Congenital Heart Defect
PDA: Persistent Ductus Arteriosus
PS: Pulmonary Stenosis
UHI: Uganda Heart Institute
UNICEF: United Nations International Children’s Emergency Fund
VSD: Ventricular Septal Defect
WHO: World Health Organisation.

## Declarations

### Data Availability

The datasets used and/or analyzed during the current study are not publicly available due to ethical and confidentiality restrictions but are available from the corresponding author on reasonable request and with permission from the Uganda Heart Institute.

### Ethical Approval and Consent to Participate

This study utilized secondary data and was conducted following approval from the Ethics Committee of the School of Statistics and Planning at Makerere University. Permission to access clinical data was obtained from the Uganda Heart Institute (UHI), which is responsible for maintaining the patient records used in this study. The original data collection procedures, including participant recruitment and informed consent, had been reviewed and approved by the relevant institutional and regulatory bodies. Informed consent was obtained from all participants or their legal guardians during the initial data collection process. As this study involved secondary analysis of existing data, the requirement for additional informed consent was waived by the Ethics Committee of the School of Statistics and Planning at Makerere University in accordance with national guidelines. All procedures performed in this study were in accordance with the ethical standards of the responsible institutional and national research committees and with the Declaration of Helsinki (1975, as revised in 2008). Strict measures were taken to ensure confidentiality and anonymity of patient information throughout the study.

### Consent for Publication

Not applicable

### Clinical Trial Number

Not applicable

